# Replacing Protein via Enteral Nutrition in a Stepwise Approach in Critically Ill Patients: A Multicenter Randomized Controlled Trial: The REPLENISH Trial Protocol

**DOI:** 10.1101/2022.11.10.22282161

**Authors:** Yaseen M Arabi, Hasan M. Al-Dorzi, Musharaf Sadat, Dina Muharib, Haifa Algethamy, Fahad Al-Hameed, Ahmed Mady, Adnan AlGhamdi, Ghaleb. A. Al Mekhlafi, Abdulrahman A Al-Fares, Ayman Kharaba, Ali Al Bshabshe, Khalid Maghrabi, Khalid Al Ghamdi, Ghulam Rasool, Jamal Chalabi, Haifaa Ibrahim AlHumedi, Maram Hasan Sakkijha, Norah Khalid Alamrey, Rabeah Hamad Alhutail, Kaouthar Sifaoui, Mohammed Almaani, Rakan Alqahtani, Ahmad S Qureshi, Mohammed Moneer Hejazi, Hatim Arishi, Samah AlQahtani, Amro Mohamed Ghazi, Saleh T Baaziz, Abeer Othman Azhar, Sara Fahad Alabbas, Mohammed AlAqeely, Ohoud AlOrabi, Alia Al-Mutawa, Maha AlOtaibi, Omar Aldibaasi, Jesna Jose, Joel Starkopf, Jean-Charles Preiser, Anders Perner, Abdulaziz Al-Dawood, the Saudi Critical Care Trials Group

## Abstract

**Background:** Protein intake is recommended in critically ill patients to mitigate the negative effects of critical illness-induced catabolism and muscle wasting. However, the optimal dose of enteral protein remains unknown. We hypothesize that supplemental enteral protein (1.2 g/kg/day) added to standard enteral nutrition formula to achieve high amount of enteral protein (range 2-2.4 g/kg/day) given from ICU day 5 until ICU discharge or ICU day 90 as compared to no supplemental enteral protein to achieve moderate amount enteral protein (0.8-1.2 g/kg/day) would reduce all-cause 90-day mortality in adult critically ill mechanically ventilated patients.

**Methods:** The REPLENISH (<u>Repl</u>acing Protein Via <u>E</u>nteral <u>N</u>utrition in a <u>S</u>tepwise Approac<u>h</u> in Critically Ill Patients) trial is an open-label, multicenter randomized clinical trial. Patients will be randomized to the Supplemental protein group or the Control group. Patients in both groups will receive the primary enteral formula as per the treating team, which includes a maximum protein 1.2 g/kg/day. The Supplemental protein group will receive, in addition, supplemental protein at 1.2 g/kg/day starting the fifth ICU day. The Control group will receive the primary formula without supplemental protein. The primary outcome is 90-day all-cause mortality. Other outcomes include functional and quality of life assessments at 90 days. The trial will enroll 2502 patients.

**Discussion:** The study has been initiated in September 2021. Interim analysis is planned at one third and two thirds of the target sample size. The study is expected to be completed by the end of 2024

**Trial Registration:** ClinicalTrials.gov Identifier: NCT04475666. Registered on July 17, 2020 https://clinicaltrials.gov/ct2/show/NCT04475666

## Background

During the acute phase of critical illness, amino acids are mobilized into the circulation in response to stress hormones to be used in tissue repair and synthesis of acute-phase proteins and other inflammatory mediators.^1, 2^ The resulting protein catabolism may be associated with immunosuppression,^3^ poor wound healing,^4^ and ICU-acquired weakness, which are associated with increased mortality and delayed recovery.^5^ Higher protein intake has been thought to mitigate the negative protein catabolic state by increasing the availability of exogenous amino acids. Consequently, clinical practice guidelines have generally recommended the administration of higher protein intake in critically ill patients than in healthy individuals (World Health Organization recommendations: 0.7-0.8 g/kg/day);^6^ however, the supportive data are limited. Observational studies showed inconsistent association between protein intake and outcomes in critically ill patients, with some studies showing that more protein was associated with better outcomes,^7-13^ others with worse outcomes,^14-16^ and others with no difference in outcomes.^17^ There is scarce evidence from randomized clinical trials (RCTs) that compared higher versus lower protein doses in ICU patients.^18-21^ A meta-analysis of 5 RCTs showed no difference in mortality with the use of higher compared to lower protein intake.^22^ The inconsistent evidence has been reflected in the variable protein doses recommended in clinical practice guidelines.^23, 24^

Additionally, the optimal timing of higher protein intake is unknown. Because protein breakdown is more pronounced in the early phase of illness, it has been suggested that higher protein intake should be given early.^13^ On the other hand, there are data suggesting that higher protein intake in the early phase of critical illness may cause harm, which may be related to inhibition of autophagy and increased ureagenesis, leading to greater muscle wasting, and delayed recovery.^25-28^

With the current state of evidence, the optimal amount of protein intake in critically ill patients remains largely unclear and is considered a high priority for research.^29-32^ The objective of this multicenter RCT is to evaluate whether supplemental enteral protein (1.2 g/kg/day) added to standard enteral nutrition formula to achieve high amount of enteral protein (range 2-2.4 g/kg/day) given from ICU day 5 or until ICU discharge up to ICU day 90 as compared with no supplemental enteral protein to achieve moderate amount enteral protein (0.8-1.2 g/kg/day) will reduce all-cause 90-day mortality in adult critically ill patients.

## Methods

### Study design and setting

The REPLENISH (<u>Repl</u>acing Protein Via <u>E</u>nteral <u>N</u>utrition in a <u>S</u>tepwise Approac<u>h</u> in Critically Ill Patients) trial is an open-label, multicenter RCT that is conducted in ICUs in Saudi Arabia and Kuwait. The study has been approved by the Institutional Review Boards of all the participating sites and sponsored by King Abdullah International Medical Research Center, Riyadh Saudi Arabia (RC19/414/R). It has been registered at ClinicalTrials.gov (NCT04475666).

SPIRIT checklist was used when writing this protocol and is attached as a supplementary file.

### Study population

All the patients will be screened for the eligibility criteria (Table 1) on ICU calendar day 4, up to the morning of ICU calendar day 5. The ICU admission calendar day is considered ICU day 1.

**Table 1:**
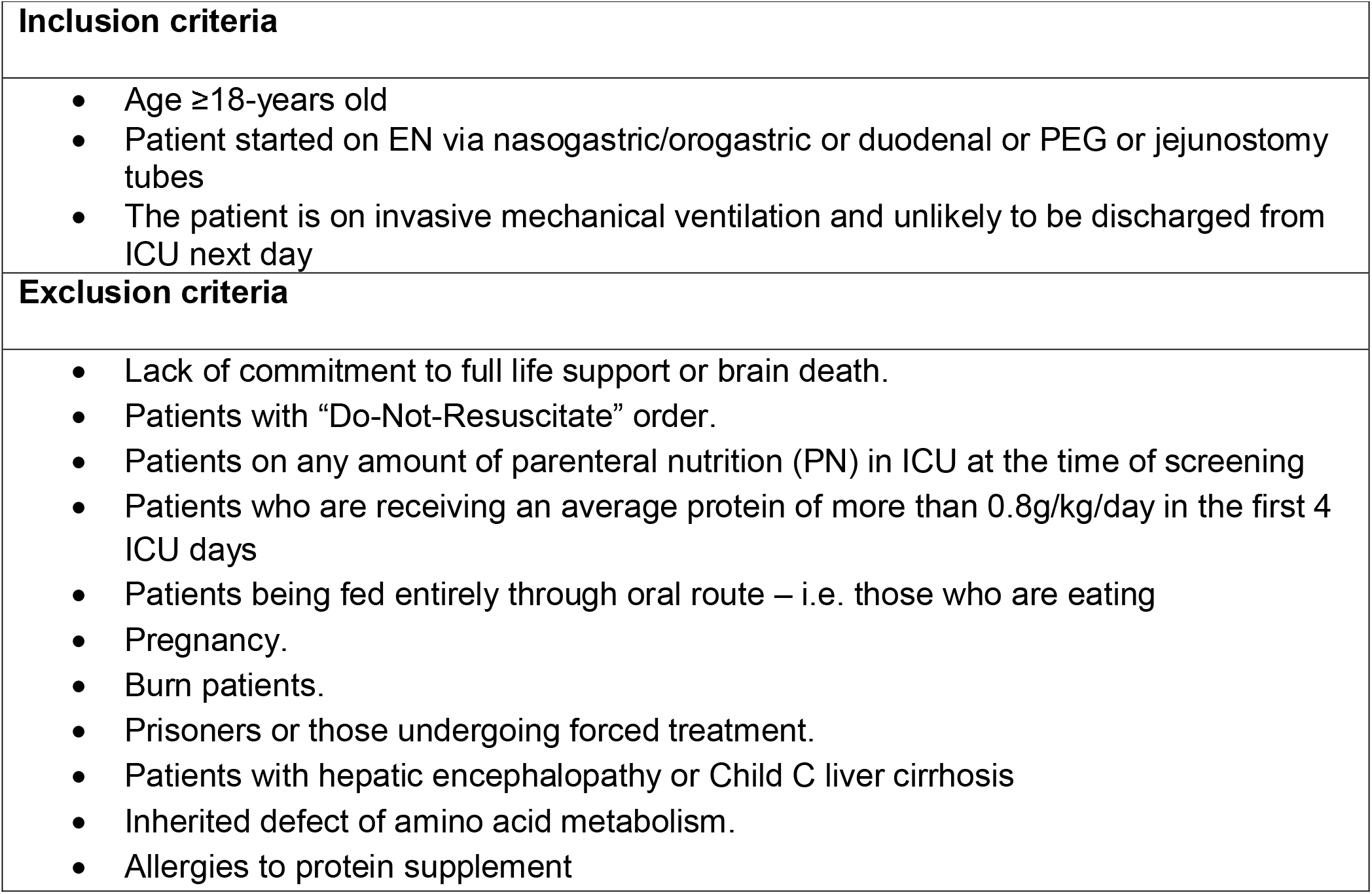
Eligibility criteria for the study.

|  |
| --- |
| **Inclusion criteria** |
| • Age ≥18-years old<br>• Patient started on EN via nasogastric/orogastric or duodenal or PEG or jejunostomy tubes<br>• The patient is on invasive mechanical ventilation and unlikely to be discharged from ICU next day |
| **Exclusion criteria** |
| • Lack of commitment to full life support or brain death.<br>• Patients with “Do-Not-Resuscitate” order.<br>• Patients on any amount of parenteral nutrition (PN) in ICU at the time of screening<br>• Patients who are receiving an average protein of more than 0.8g/kg/day in the first 4 ICU days<br>• Patients being fed entirely through oral route – i.e. those who are eating<br>• Pregnancy.<br>• Burn patients.<br>• Prisoners or those undergoing forced treatment.<br>• Patients with hepatic encephalopathy or Child C liver cirrhosis<br>• Inherited defect of amino acid metabolism.<br>• Allergies to protein supplement |

### Inclusion criteria

1. Age ≥18-years old.
2. The patient is started on enteral nutrition via feeding tube (naso/oro-gastric, naso/oro-enteral, gastrostomy or jejunostomy tubes).
3. The patient is on invasive mechanical ventilation and unlikely to be discharged from ICU next day.

### Exclusion criteria

1. Lack of commitment to full life support or brain death. Patients with “Do-Not-Resuscitate” order but with commitment to ongoing life support can be enrolled.
2. The patient is on any amount of parenteral nutrition (PN) in ICU at the time of screening, whether PN is used alone or in combination with enteral nutrition. Non-nutritional calories (dextrose, propofol, citrate) not considered as PN.
3. The patient has received an average protein of more than 0.8 g/kg/day in the first 4 ICU days.
4. The patient is fed entirely through oral route – i.e., those who are eating.
5. The patient has hepatic encephalopathy or Child C liver cirrhosis
6. The patient is admitted because of burn.
7. The patient has an inherited defect of amino acid metabolism.
8. The patient has allergy to protein supplement.
9. Pregnancy.
10. Prisoners or those undergoing forced treatment.

### Recruitment

The research team will approach the patient or surrogate decision maker for consent according to local regulations. Because both feeding strategies are within the standard of care and because enrollment needs to be done early to initiate the feeding strategy, deferred consent can be used if consent could not be obtained a priori. The research coordinator will be maintaining a screening log of eligible patients who are not randomized.

### Randomization

Enrolled patients will be randomized through a web-based system at a 1:1 ratio to Supplemental protein group or Control group using permuted variable undisclosed block sizes. Randomization will be stratified by the trial site, the use of renal replacement therapy at the time of randomization and whether the patient is a suspected or confirmed case of COVID-19.

### Coenrollment

Co-enrollment in other RCTs is permitted after approval by both trial steering committees.

### Nutrition in the two study groups

#### Energy in both groups

Until ICU calendar day 4, the prescription of energy will be left to the discretion of the treating teams. If desired, energy expenditure can be determined using the predictive equations or indirect calorimetry, based on the practice at individual sites. Between days 5 and 90, the energy target is 70 to 100% of calculated or measured caloric requirements. Caloric intake will be calculated taking into consideration intravenous dextrose, citrate and propofol. Caloric intake will include the administered protein in the primary formula.

#### Protein Pre-randomization (ICU calendar day 1-4)

Until ICU day 4, protein requirement will be provided according to the local practice as long as no intravenous amino acids are given and the average protein intake in the first 4 days does not exceed 0.8 g/kg/day.

#### Protein Post-randomization (ICU day 5-ICU discharge) in the Control group

The subjects randomized to the Control group will receive standard prescription without supplemental proteins (maximum1.2 g/kg/day) from the primary polymeric formula. No supplemental protein will be allowed. For patients with BMI <30 kg/m^2^, we will use pre-ICU actual body weight and if unavailable the weight on ICU admission. For patients with BMI ≥30 kg/m^2^, we will use adjusted body weight (Table 2).^24^

**Table 2:**
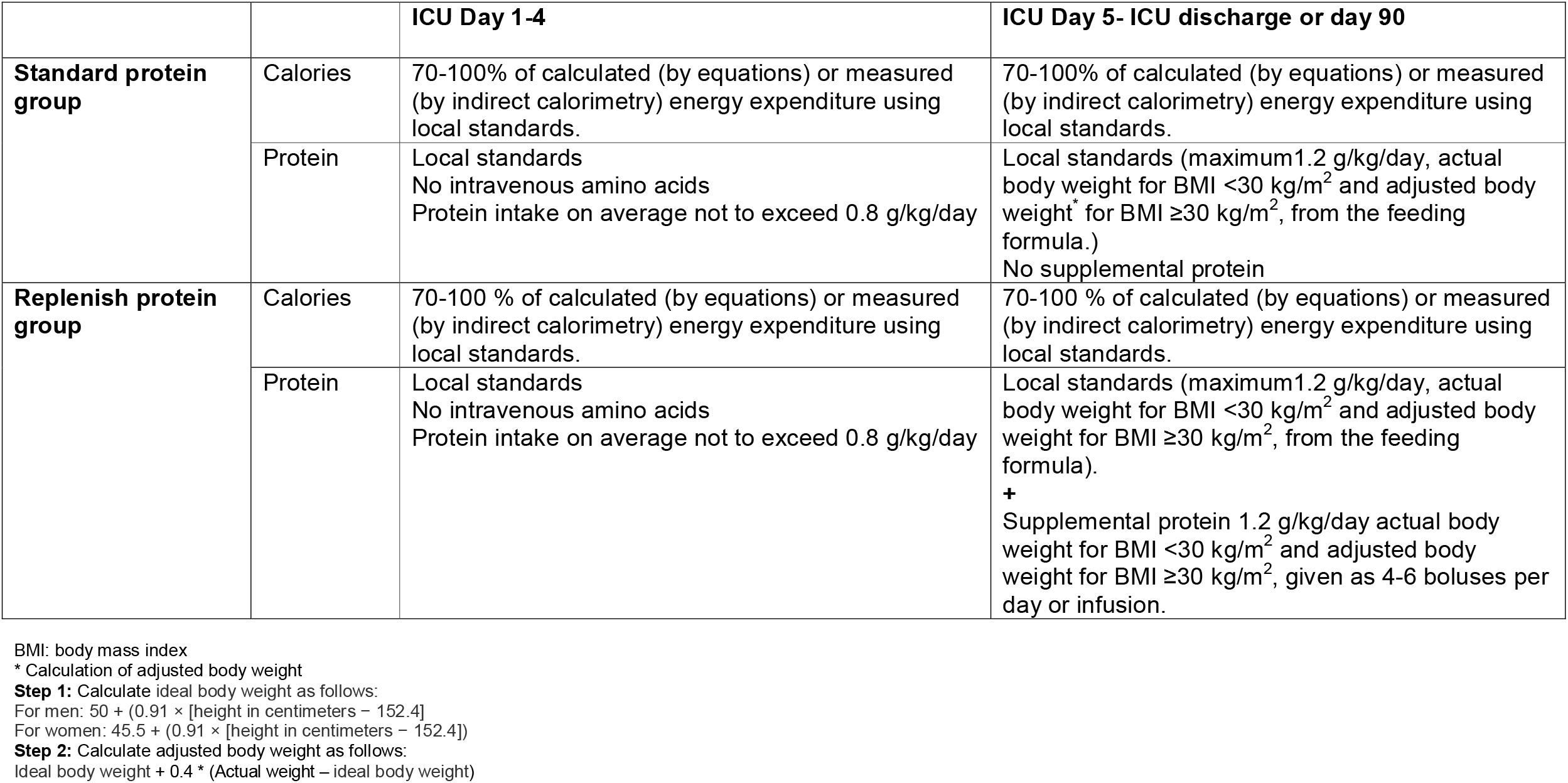
Daily protein provision for patients enrolled in the trial.

|  |  | **ICU Day 1-4** | **ICU Day 5- ICU discharge or day 90** |
| --- | --- | --- | --- |
| **Standard protein group** | Calories | 70-100% of calculated (by equations) or measured (by indirect calorimetry) energy expenditure using local standards. | 70-100% of calculated (by equations) or measured (by indirect calorimetry) energy expenditure using local standards. |
|  | Protein | Local standards<br>No intravenous amino acids<br>Protein intake on average not to exceed 0.8 g/kg/day | Local standards (maximum 1.2 g/kg/day, actual body weight for BMI <30 kg/m<sup>2</sup> and adjusted body weight* for BMI ≥30 kg/m<sup>2</sup>, from the feeding formula.)<br>No supplemental protein |
| **Replenish protein group** | Calories | 70-100 % of calculated (by equations) or measured (by indirect calorimetry) energy expenditure using local standards. | 70-100 % of calculated (by equations) or measured (by indirect calorimetry) energy expenditure using local standards. |
|  | Protein | Local standards<br>No intravenous amino acids<br>Protein intake on average not to exceed 0.8 g/kg/day | Local standards (maximum 1.2 g/kg/day, actual body weight for BMI <30 kg/m<sup>2</sup> and adjusted body weight for BMI ≥30 kg/m<sup>2</sup>, from the feeding formula).<br>**+**<br>Supplemental protein 1.2 g/kg/day actual body weight for BMI <30 kg/m<sup>2</sup> and adjusted body weight for BMI ≥30 kg/m<sup>2</sup>, given as 4-6 boluses per day or infusion. |

BMI: body mass index
\* Calculation of adjusted body weight
**Step 1:** Calculate ideal body weight as follows:
For men: $50 + (0.91 \times [\text{height in centimeters} - 152.4])$
For women: $45.5 + (0.91 \times [\text{height in centimeters} - 152.4])$
**Step 2:** Calculate adjusted body weight as follows:
Ideal body weight + $0.4 \times (\text{Actual weight} - \text{ideal body weight})$

#### Protein Post-randomization (ICU day 5 to ICU discharge) in the Supplemental protein group

The subjects randomized to the Supplemental protein group will receive the standard amount of proteins (maximum 1.2 g/kg/day) from the primary formula with the addition of supplemental enteral protein at 1.2 g/kg/day. Supplemental protein is administered as boluses by syringe through the feeding tube with flushes with a minimum of 30-60 ml of water. The choice of supplemental protein is left to the local teams as per availability.

### Co-interventions

a. PN: Patients on any amount of PN at the time of screening will not be enrolled in the trial. However, patients who have been enrolled in the trial and deemed to need PN by their treating team, will remain in the trial.
b. Glucose management: All centers may use their own standard protocols as long as the target blood glucose is between 4.4-10 mmol/L (80-180 mg/dl).
c. Mobility assessment: All centers may use their own standard protocols with regards to mobility in ICU patients. Data on mobility will be collected.
d. Medications: Data on corticosteroids and statins will be collected during the ICU stay (Up to day 90)
e. Selection of enteral nutritional formula: The type of formula will be left to the discretion of the attending physician. The types of formula used are grouped as general or disease non-specific (for example: Osmolite, Resourse, Resourse plus, Ensure, Ensure plus, Jevity 1.0 and Jevity 1.2) or disease specific (For example: Pulmocare, Glucerna, Suplena, Peptamen 1.0, Peptamen 1.2, Peptamen 1.5, Novasource Renal, Nepro, Nutren hepatic, Promote and Vivonex plus).
f. The enteral feeding protocol: Each ICU may use their own enteral feeding protocol. The use of prokinetics and type of feeding tube (large-bore nasogastric tube or small-bore nasogastric tube with or without guide wire) is left to the treating team.
g. Multivitamins: Patients included in the trial in both arms will receive enteral multivitamins as per local formulary.

### Duration of the intervention

The study intervention will continue until meeting any of the following criteria: death, ICU discharge or day 90 in ICU, premature stopping of feeding due to brain death or palliative care plan whichever comes first, initiation and tolerating of full oral feeding for more than 24 hours (i.e., treating physicians feel that enteral nutrition is no longer required). In these situations, the study intervention will no longer be followed, and nutrition will be at the discretion of the treating teams but outcome data will be collected. Figure 1 shows the schedule of enrollment, intervention and assessment for the trial according to the SPIRIT template.

**Figure 1:**
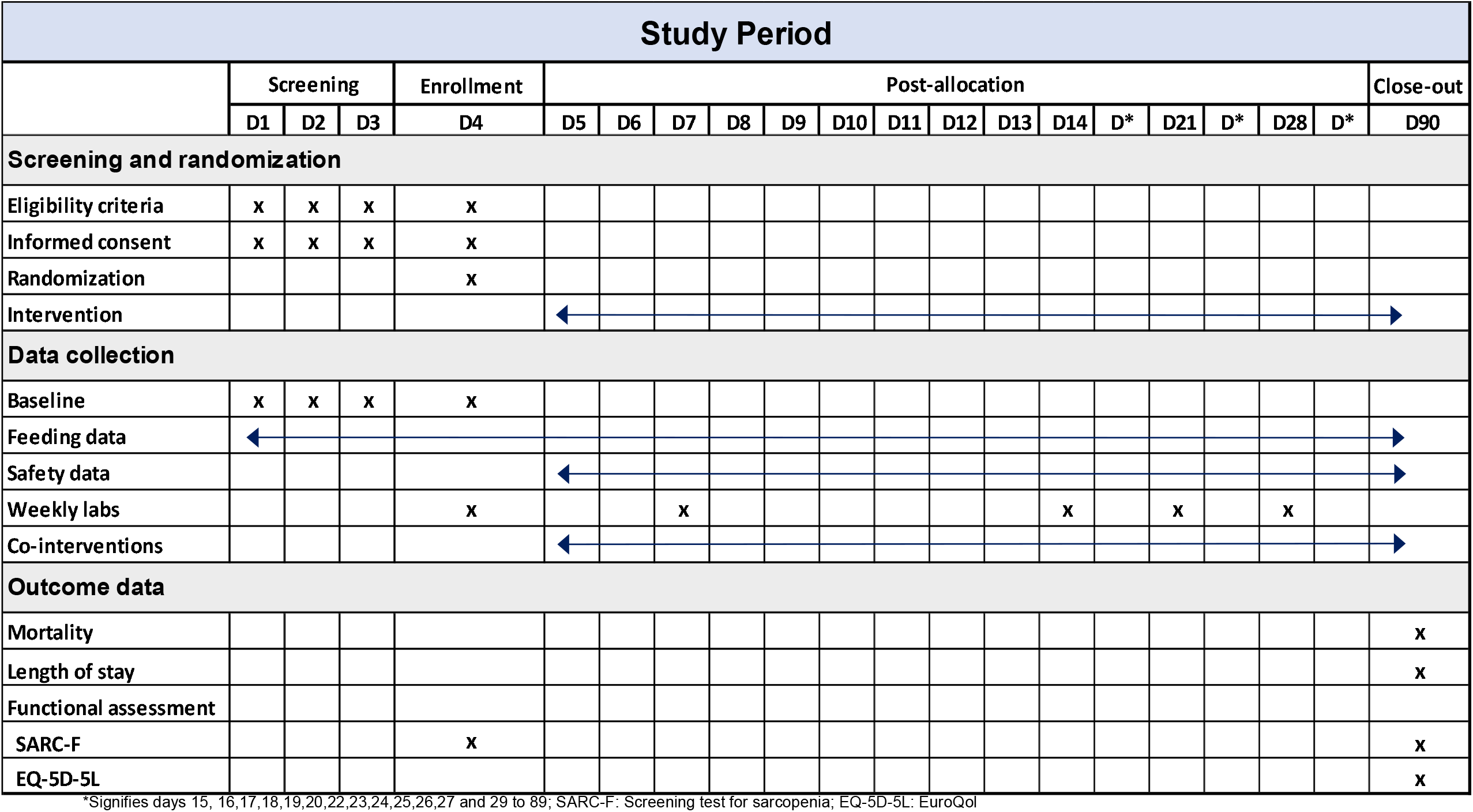
Timeline for screening, enrollment and assessment for patients enrolled in REPLENISH trial.

### Data collection

Baseline data collected from ICU days 1 to 4 will include age, sex, admission category (medical, postoperative (non-trauma) and trauma (post-operative and non-operative)), comorbidities (defined as per the Acute Physiology and Chronic Health Evaluation (APACHE) II system), APACHE II score, simplified mortality score (SMS),^33^ Sequential Organ Failure Assessment (SOFA) score on day 4 and, pre-morbid functional assessment using SARC-F screen for sarcopenia.^34^ For patients with COVID-19, additional baseline labs including ferritin, interleukin-6, lactate and procalcitonin will be collected. Daily data collected from ICU days 1 to 90 or ICU discharge will include nutritional data including energy and protein administered daily, blood glucose and insulin data, vasopressor use, use of renal replacement therapy, use of invasive mechanical ventilation, creatinine, blood urea nitrogen and urine output. Additional data collected on days 4, 7, 14, 21, 28 will include mobility level assessment^35^, optional labs for selected sites only (prealbumin, albumin, ammonia, 24-hour urine for urinary urea nitrogen, lowest potassium level, lowest magnesium level, lowest phosphate level, aspartate transaminase, alanine aminotransferase and international normalized ratio). In case the labs have multiple readings in a day, the worst values will be recorded.

### Outcome measures

The primary outcome is 90-day all-cause mortality. Secondary outcomes are days alive at day 90 without life support (use of vasopressor/inotropic support, invasive mechanical ventilation and/or renal replacement therapy), days alive and out of the hospital at day 90, bacteremia until 2 days post ICU, new or progression of skin sacral pressure ulcers in ICU,^36^ functional assessment using SARC-F screen for sarcopenia,^34^ and EuroQoL (EQ)-5D-5L) at day 90. Safety outcomes are classified into major and minor safety outcomes. Major safety outcomes are new episode of stage 2 or higher acute kidney injury by KDIGO criteria^37^ after enrollment, newly confirmed pneumonia according to the modified CDC criteria^38^, grade IV acute gastrointestinal injury,^39^ including any of bowel ischemia with necrosis, clinically important gastrointestinal bleeding, Ogilvie’s syndrome, and abdominal compartment syndrome. Additionally, minor safety outcomes will be recorded as one or more of the following: feeding intolerance, diarrhea,^39^ and refeeding syndrome^40, 41^ (Supplementary Table).

### Trial management

a. ***Steering committee:*** The study Steering Committee will be responsible for overseeing the management of the trial, providing training and support to participating centers for protocol adherence, upholding or modifying study procedures as needed and the statistical analysis plan. This will be achieved through virtual or in-person meetings every 3-6 months.
b. ***Ethics approval:*** The study has been approved by the Institutional Review Board of the Ministry of National Guard Health Affairs, Riyadh, Saudi Arabia. All participating sites will obtain approval from the related Institutional Review Boards. The study will be conducted in accordance with the ethical principle of the Declaration of Helsinki and International Council for Harmonization–Good Clinical Practice guidelines.
c. ***Data management:*** Patient data will be de-identified and stored in a secure server at King Abdullah International Medical Research Center, Riyadh. The database will be password protected and the participating sites will have a unique credentials to access the database.
d. ***Protocol compliance:*** Several measures will be taken to ensure optimal compliance with the study protocols. Before launching the study, ICU physicians, nurses and dietitians will attend training sessions, with special emphasis on achievement of the protein target as per protocol. Follow-up training sessions will be conducted periodically to provide feedback. The adherence to the protocol and data quality will be monitored by the coordinating site on regular basis, at least every three months. Feedback will be provided to each site to further improve adherence to nutritional targets.
e. ***Loss-to-follow up:*** Patients will be followed post ICU discharge (without further intervention) to document hospital vital status. However, if the randomized patient at any point decides to withdraw from the trial intervention at the request of either the patient himself, family, or the treating physician, the data will be included in the group to which they were allocated as per the intention-to-treat principle and the reason of withdrawal will be documented.
f. ***90-day follow-up:*** 90-day outcomes will be documented from the chart or registries or, if needed, by a telephonic interview from the patient or next of kin if the patient is discharged alive. 90-day follow up will include vital status, date of death if the patient is dead and functional assessment using SARC-F screen and EuroQoL (EQ)-5D-5L) if the patient is alive on that day.
g. ***Safety Monitoring:*** Serious adverse events that are suspected related to research procedures will be reported as per local guidelines. However, the trial involves a low-risk intervention, and the two levels of protein intake under study (high versus low) are within the recommended ranges of administered protein supplementation to ICU patients by most Clinical Practice Guidelines. Therefore, it is anticipated that most of adverse events occur as part of the participants’ natural disease process. Safety outcomes as well as serious adverse events will be reported to the Data Safety Monitoring Board.

### Statistical methods

a. ***Sample size*:** We anticipate a baseline 90-day mortality of 30% and an absolute risk reduction of 5% with the high-protein intervention. The baseline risk was estimated based on a similar cohort from the Permissive Underfeeding or Standard Enteral Feeding in Critically Ill Adults (PermiT trial) and Pantoprazole in patients at risk for gastrointestinal bleeding in the ICU (SUP-ICU) trials. In the PermiT trial which included patients from 7 sites in Saudi Arabia and Canada,^42^ 715 patients received mechanical ventilation for >4 days, and 209 died by day 90 (29.3%). In the SUP-ICU trial,^43^ 48% (1571/3282) of all included patients were mechanically ventilated on day 4.Of these, 34% (530/1571) had died on day 90. The SUP-ICU trial enrolled acutely admitted ICU patients with at least one risk factor for GI bleeding in 33 ICUs in 5 countries in Northern Europe. The treatment effect in REPLENISH (5% absolute risk reduction) was based on a propensity-score adjusted analysis which showed an odds ratio for the association of high protein compared to a moderate protein of 0.80 (95% CI 0.56, 1.16, p=0.24)^17^. The final analysis of the primary outcome will be based on two-sided alpha (α) of 0.05 and power (1-β) = 0.80. Based on these assumptions, we need 1251 patients in each group (2502 in both groups).
b. ***Statistical analysis*:** The analyses will be done in the intention-to-treat population defined as all randomized patients for whom there is consent for the use of data. Baseline characteristics will be summarized as numbers and percentages (categorical variables), and continuous variables will be summarized as medians and first and third quartiles (Q1, Q3) or means and standard deviation. We will compare the proportions for the primary and secondary outcomes between patients randomized to standard protein versus high-protein group. We will calculate the relative risk reduction, absolute risk reduction, and the number needed to treat to prevent one death. We will present the primary result with 95% confidence intervals and a 2-sided p-value (5% level of significance). A detailed statistical analysis plan will be developed and published before trial completion. Each component of the composite outcomes, such as serious adverse events and use of life support, will be reported in a supplement to the primary publication, but any differences between these single components will not be analysed.
c. ***Subgroup analyses*:** The following prespecified subgroups will be analyzed based on admission category: Medical vs postoperative vs trauma patients. In addition, we will evaluate the effect of the intervention within subpopulations of our enrolled patients: day 4 SOFA stratified at a value of 9, admission diagnosis of sepsis versus others, severe traumatic brain injury versus others, vasopressor use at the time of enrollment versus none, acute kidney injury at enrollment (4 KDIGO groups: 0, 1, 2, 3) and COVID-19 status.
d. **Interim analysis:** Interim analysis is planned when 33% and 67% of the sample size has been achieved. The trial may be stopped for safety (based on mortality) (p<0.01) or effectiveness (<0.001) or if there is other compelling evidence that trial participants are being harmed. There will be no plans to terminate the trial for futility. We will account for alpha spending by the O’Brien Fleming method and the final significance level will be 0.048.^44^

### Sub-Studies

a. ***REPLENISH-COVID sub-study:*** In this sub-study we will evaluate the effect on high-protein on the subgroup of COVID-19 patients.
b. ***Quality of life:*** A detailed analysis of the effect of protein on quality of life will be reported.

## Discussion

In this REPLENISH trial, we hypothesize that supplemental enteral protein starting the fifth ICU day in addition to standard enteral nutrition will improve the survival of adult critically ill patients compared to standard enteral nutrition alone. The results of the trial address an important question in enteral nutrition in critically patients.

The study design took in consideration multiple factors as learnt from a pilot study (the REPLENISH Pilot trial).^45^ In the current study, we followed a pragmatic approach to eligibility criteria, management of protein and energy intake and selection of outcome measurements.^45^ For example, with the exception of protein intake, all other aspects of nutrition management were left to the treating teams. The administration of protein as boluses instead of infusion facilitates a closer achievement of protein targets.

### Trial status

The trial started in September 2020 and has recruited 711 patients from 14 sites in Saudi Arabia and one in Kuwait.. The study was started according to the protocol in its third version dated June 03, 2020. There have been minor protocol modifications after starting the trial (versions 4 and 5). These changes have no impact on the study conduct. The trial is expected to complete patient follow-up by the end of 2024. Upon completion, the results of the trial will be submitted to a peer-reviewed journal. Authorship will be guided by the Uniform Requirements for Manuscripts Submitted to Biomedical Journals (URM) developed by ICMJE

## Data Availability

The data that will support the findings of this study are available from the corresponding author upon reasonable request as per the regulations of King Abdullah International Medical Research Center (KAIMRC)

## List of abbreviations

APACHE: Acute Physiology and Chronic Health Evaluation
DSMB: Data Safety Monitoring Board
ESPEN: European Society for Clinical Nutrition and Metabolism
ICU: Intensive Care Unit
KDIGO: Kidney Disease: Improving Global Outcomes
PN: Parenteral Nutrition
RCT: Randomized Controlled Trial
SOFA: Sequential Organ Failure Assessment

## Declaration

### Ethics approval and consent to participate

The study protocol as well as the informed consent have been approved by the National Guard Health Affairs Institutional Review Board (RC19/414/R), Riyadh and the respective Institutional Review Boards of all the other centers.

### Consent for publication

Not applicable

## Competing interests

The authors declare that they have no competing interests.

### Funding

The study is sponsored by King Abdullah International Medical Research Center, Riyadh, Saudi Arabia.

### Author contributions

YA is the Chief Investigator; conception, design and development of the protocol, analytical plan, drafting and critical revision of the manuscript for important and intellectual content. HMD, MS, DM, HG, FH, AM, AG, GM, AF, AK, AB, KM, KG, GR, JC, HIA, MHS, NA, RH, KS, MA, RQ, AQ, MH, HA, SQ, AG, SB, AO, SF, MA, OA, AAM, MO, OD, JJ, JS, JCP, AP and AD contributed to the development of the protocol, critical revision of the manuscript for important intellectual content. All authors read and approved the final manuscript.

## Acknowledgement

We would like to thank the Data Safety Monitoring Board Chair and members:

## Michelle Ng Gong, M.D, M.S.(Chair)

Professor of Medicine

Professor of Epidemiology and Population Health

Albert Einstein College of Medicine

Chief, Department of Medicine Division of Critical Care

Chief, Department of Medicine Division of Pulmonary Medicine

Director, Department of Medicine Critical Care Research

Montefiore Medical Center, Bronx, NY, United States of America

### Manpreet Singh Mundi, MD (Member)

Professor, Department of Medicine,

Division of Endocrinology, Diabetes, Metabolism, and Nutrition

Mayo Clinic, Rochester, MN

United States of America

### Christopher John Lindsell (Member)

Professor of Biostatistics and Biomedical Informatics

Director, Vanderbilt Institute for Clinical and Translational Research (VICTR) Methods Program

Co-director, Vanderbilt Health Data Science (HEADS) Center

Nashville Tennessee

United States of America

## Collaborators

### The Saudi Critical Care Trials Group

Amal Almatroud, Brintha Naidu, Vicki Burrow, Salha Al Zayer, Haseena Banu Khan, Afonso Varela,

Mohamed Ali Alodat, Rayan Alshayeh, AbdulRehman AlHarthi, Naif Al Qahtani, Yasmeen Ayed AlHejiely, Mada Muzhir AlZahrani, Mohammed Haddad Lhmdi, Katrina Baguisa, Huda Mhawisg,

Liyakat Khan, Moataz Gabr, Shehla Nuzhat, Madiha ElGhannam, Beverly Bcuizon, Bander AlAnezi, Christine Joy Anaud, Sawsan Albalawi, Manar Alahmadi, Mohammed AlHumaid, Samar Talal Nouri, Rozeena Huma, Khawla Farhan, Samahar Alamoudi, Milyn L Ansing, Raghad Malabari, Kholoud Shobragi, Shaymaa Asaas, Ahmed Quadri, Khalid Idrees, Arwa AlHusseini, Shahinaz Bashir, Mohamed Hussein, Olfa Baji, Abdulrehman Alerw, Khloud Johani, Monera AlEnezi, Ismail Boudrar, Rabiah Atiq, Maali Junid, Maram Yusef, Mona Bin Mabkoot, Munir AlDammad, Yahia Otaif, Osama Hakami, Mariam Ehab Kenawy, Dalal Ali Alkhamees, Tasneem Abdullah Behbehani

