## supplementary file for "Replacing Protein via Enteral Nutrition in a Stepwise Approach in Critically Ill Patients: A Multicenter Randomized Controlled Trial: The REPLENISH Trial Protocol"

| **Table 1:** REPLENISH Trial management, Steering committee, Data safety monitoring board members and Site collaborators. | |
| --- | --- |
| **Management Committee** | Yaseen M. Arabi  Hasan M. Al-Dorzi  Abdulaziz Al-Dawood  Musharaf Sadat  Omar Al Dibaasi  Jesna Jose  Haifa AlHumedi |
| **Writing Committee** | Yaseen M. Arabi  Hasan M. Al-Dorzi  Musharaf Sadat  Omar Al Dibaasi  Jesna Jose |
| **Data Safety Monitoring Board** | Michelle Ng Gong  Manpreet Singh Mundi  Christopher John Lindsell |
| **Collaborators - the Saudi Critical Care Trials Group** |  |
| **King Saud bin Abdulaziz University for Health Sciences and**  **King Abdullah International Medical Research Center, Riyadh, Saudi Arabia** | Yaseen M. Arabi  Hasan M. Al-Dorzi  Abdulaziz Al-Dawood  Omar Al Dibaasi  Jesna Jose  Musharaf Sadat  Haifa AlHumedi  Amal Almatroud  Brintha Naidu  Vicki Burrow  Salha Al Zayer  Haseena Banu Khan  Afonso Varela  Maram Sakkijha  Norah Khalid Alamrey  Hatim Areshi  Samah Qahtani  Amro Ghazi  Mohammed Moneer Hejazy |
| **King Saud Medical City, Riyadh** | Dina Muharib  Ahmed Mady  Mohammed AlAqeely  Maha AlOtaibi  Mohamed Ali Alodat  Rayan Alshayeh  AbdulRehman AlHarthi  Naif Al Qahtani  Yasmeen Ayed AlHejiely  Mada Muzhir AlZahrani  Mohammed Haddad Lhmdi  Katrina Baguisa  Huda Mhawisg |
| **King Abdulaziz university hospital, Jeddah** | Haifa AlGhethamy  Saleh T Baaziz  Abeer Othman Azhar  Sara Fahad Alabbas  Liyakat Khan  Moataz Gabr  Shehla Nuzhat |
| **King Fahad Medical City, Riyadh, Saudi Arabia** | Mohammed AlMaani |
| **Prince Sultan Military Medical City, Riyadh, Saudi Arabia** | Adnan AlGhamdi  Ghaleb A. AlMekhlafi  Rabeah Hamad Alhutail  Madiha ElGhannam  Beverly Bcuizon  Bander AlAnezi  Christine Joy Anaud |
| **King Faisal Specialist Hospital & Research Center, Riyadh, Saudi Arabia** | Khalid Maghrabi  Sawsan Albalawi  Manar Alahmadi  Mohammed AlHumaid  Samar Talal Nouri  Rozeena Huma  Khawla Farhan |
| **King Faisal Specialist Hospital & Research Center, Jeddah, Saudi Arabia** | Khalid Al Ghamdi  Lama Hefni  Samahar Alamoudi  Milyn L Ansing |
| **King Abdulaziz Medical City, Jeddah, Saudi Arabia** | Fahad Al-Hameed  Ohoud Aloraabi  Ghulam Rasool  Raghad Malabari  Kholoud Shobragi  Shaymaa Asaas |
| **Prince Mohammed bin Abdulaziz Hospital, Madinah, Saudi Arabia** | Ahmad S Qureshi  Ahmed Quadri  Khalid Idrees |
| **King Abdulaziz Hospital-Alahsa, Saudi Arabia** | Jamal Chalabi  Arwa AlHusseini  Shahinaz Bashir |
| **Ohoud Hospital, Al-Madinah Al-Monawarah, Saudi Arabia** | Ayman Kharaba  Mohamed Hussein  Kaouthar Sifaoui  Olfa Baji  Abdulrehman Alerw  Khloud Johani  Monera AlEnezi  Ismail Boudrar  Rabiah Atiq  Maali Junid  Maram Yusef |
| **Aseer Central Hospital, Abha, Saudi Arabia** | Ali Al Bshabshe  Munir AlDammad  Yahia Otaif  Osama Hakami |
| **Al-Amiri Hospital, Ministry of Health, Kuwait** | Abdulrahman A Al-Fares  Alia Al-Mutawa |
| **King Saud University, Riyadh Saudi Arabia** | Rakan Alqahtani  Mona Bin Mabkoot |
| **Erasme University Hospital, Brussels, Belgium** | Jean-Charles Preiser |
| **University of Tartu, Estonia** | Joel Starkopf |
| **University of Copenhagen, Denmark** | Anders Perner |

| **Table 2:** Study outcome and their definitions. | | |
| --- | --- | --- |
| **Variables** | **Outcomes** | **Definitions** |
| **Primary outcome** | 90-day all-cause mortality | Death within 90 days from ICU admission |
| **Secondary outcomes** | Days alive at day 90 without life support | Without use of vasopressor/inotropic support, invasive mechanical ventilation and/or renal replacement therapy) |
|  | Days alive and out of hospital at day 90 |  |
|  | Bacteremia | Positive blood cultures until 2 days post ICU |
|  | New or progression of skin pressure ulcers in sacral areas | Skin assessment staging  Stage I: Non-blanchable erythema  Stage II: Partial thickness  Stage III: Full thickness skin loss  Stage IV: Full thickness tissue loss. |
|  | Day 90 Functional assessment | SARC-F screen for sarcopenia[12] |
|  | Day 90 functional assessment | Evaluate by EuroQoL (EQ)-5D-5L) |
| **Safety outcomes** | | |
| **Major safety outcomes** | New episode of stage 2 or higher acute kidney injury by KDIGO criteria after enrollment | Stage 2: Increase in creatinine 2.0 to 2.9 multiplied by baseline serum creatinine OR urine output <0.5 ml/kg/hr for ≥ 12 hours  Stage 3: Increase in creatinine ≥ 3.0 multiplied by baseline serum creatinine OR increase to > 4.0 mg/dl (353.6 micromol/L) OR new renal replacement therapy OR urine output <0.3 ml/kg/hr for 24 hours OR anuria for > 24 hours |
|  | Pneumonia defined as episodes of newly confirmed pneumonia according to the modified CDC criteria | Two or more serial chest radiographs with at least one of the following:   - New or progressive and persistent infiltrate - Consolidation - Cavitation   AND  At least one of the following:   - Fever (>38°C) with no other recognized cause - Leukopenia (white cell count < 4 x 10^9^ /l) or leukocytosis (white cell count >12 x 10^9^ /l)   AND  At least two of the following:   - New onset of purulent sputum or change in character of sputum, or increased respiratory secretions or increased suctioning requirements - New onset or worsening cough, or dyspnea, or tachypnea - Rales or bronchial breath sounds - Worsening gas exchange (hypoxemia, increased oxygen requirement, increased ventilator demand) |
|  | Grade IV Acute Gastrointestinal injury (AGI)[13] | **Bowel ischemia** defined as any of the following:   - Absent blood flow in one of the main arteries supplying the bowel with evidence of bowel wall compromise on an imaging study (CT angiography, angiography, or magnetic resonance angiography) - Presence of endoscopy criteria for colonic ischemia according to the Favier classification system (stage I, petechiae; stage II, petechiae and superficial ulcers; and stage III, necrotic ulcers and polypoid lesions) - Evidence of bowel ischemia during surgery. |
|  |  | **Clinically important gastrointestinal bleeding** defined as overt gastrointestinal bleeding and at least one of the following four features within 24 hours of gastrointestinal bleeding (in the absence of other causes) in the intensive care unit   - Spontaneous drop of systolic blood pressure, mean arterial pressure or diastolic blood pressure of 20 mmHg or more - Start of vasopressor or a 20% increase in vasopressor dose - Decrease in hemoglobin of at least 2 g/dl (1.24 mmol/l) or - Transfusion of two units of packed red blood cells or more. |
|  |  | **Ogilvie’s syndrome** defined as bowel dilatation if colonic diameter exceeds 6 cm (greater than 9 cm for cecum) or small bowel diameter exceeds 3 cm, diagnosed either on plain abdominal X-ray or CT scan without underlying mechanical obstruction or other organic cause |
|  |  | **Abdominal compartment syndrome** is defined as a persistent intra-abdominal pressure (IAP) of more than 20 mmHg accompanied by new organ dysfunction or failure |
| **Minor safety outcomes** | Feeding intolerance | Vomiting or large gastric residual volume (GRV) ≥500 ml |
|  | Diarrhea | Three or more loose or liquid stools per day with a stool weight greater than 200–250 g/day (or greater than 250 ml/day) |
|  | Refeeding syndrome[14, 15] | Serum phosphate below 0.65 mmol/L within 72 hours of starting intervention  Or  A drop in serum phosphate by > 0.16 mmol/L from a previously recorded reading within 72 hours of starting intervention |
